# Predicting Genetic Risk for Impulsivity and Substance Use in Adolescence

**DOI:** 10.1101/2025.04.07.25325258

**Authors:** Natasha E. Wade, Jonathan Ahern, Veronica Szpak, Alexander L. Wallace, Ryan M. Sullivan, Chun C. Fan, Robert Loughnan

**Affiliations:** Department of Psychiatry, University of California, San Diego, CA, USA; Department of Cognitive Science, University of California, San Diego, 9500 Gilman Drive, La Jolla, CA 92093; Center for Population Neuroscience and Genetics, Laureate Institute for Brain Research, Tulsa, OK 74103, USA; Center for Human Development, University of California, San Diego, 9500 Gilman Drive, La Jolla, CA 92093; J. Craig Venter Institute, La Jolla, CA, USA, 4120 Capricorn Ln, La Jolla, CA 92037

## Abstract

Studying genetic contributions to substance initiation is crucial for identifying at-risk individuals and developing targeted prevention strategies. Investigating these factors during adolescence is vital, as this period is critical for brain development and represents an age of experimentation and initiation of substance use. Here we generate polygenic scores (PGSs), using data from the PGS catalog, across a range of substance use related traits to assess PGS in predicting i) measures of impulsivity taken from the UPPS-P questionnaire and ii) self-reported use of nicotine/tobacco, cannabis, alcohol and caffeine in early-mid adolescence. Repeat cross-sectional analyses across age bands (ages 9-10, 11-13, and 13-15) were conducted using the longitudinal Adolescent Brain Cognitive Development (ABCD) Study^®^ (total N=8,753; 55% female). Due to the large contribution of European-like (EUR-like) individuals in discovery samples, we performed ancestry stratified analysis in EUR-like (n=5,225), African (AFR-like; n=637) and admixed (MIX-like; n=2,891) groups reflecting genetic similarity to continental ancestry groups. In the EUR-like group, PGS related to nicotine/tobacco were associated with greater impulsivity across all subscales of the UPPS-P at all ages. Analyses across ages 9-15 years old revealed PGS-impulsivity associations that: a) grew as the sample aged (e.g. Smoking Status PGS with Lack of Perseverance: 9-10 years-old: β=0.065, 11-13 years-old: β=0.11, 13-15 years-old: β=0.12) and b) others that diminished as the sample aged (e.g. Alcohol Consumption PGS with Sensation Seeking: 9-10 years-old: β=0.070, 11-13 years-old: β=0.062, 13-15 years-old: β=0.03). Evaluating the performance of PGS against self-reported substance use, PGS of nicotine/tobacco traits were associated with regular consumption of caffeine across ages. At ages 13-15, PGS of smoking traits were associated with cannabis and tobacco exposure (e.g., Smoking Initiation PGS and self-reported cannabis use, ΔR^2^=0.0094), in addition to weekly caffeine consumption. Across ages, nicotine/tobacco and alcohol PGS and regular energy drink consumption associations grew over time (e.g., Smoking Status PGS: 9-10 years-old: β=0.088, 11-13 years-old: β=0.24, 13-15 years-old: β=0.29). As with impulsivity, some PGS associations decreased over time (Alcohol Consumption PGS and self-reported alcohol use: 9-10 years-old: β=0.12, 11-13 years-old: β=0.11, 13-15 years-old: β=0.083). Replication of our EUR-like results in AFR-like and MIX-like sub-samples revealed a significant attenuation of effects, underscoring the importance of collecting genetic studies in larger ancestrally diverse cohorts. Our results highlight the dynamic relationship between genetic risk factors of substance use, trait impulsivity, and self-reported substance initiation throughout adolescence. Further, evidence here indicates caffeine consumption represents an early risk factor for problematic substance use in later life. Results support PGSs, in conjunction with larger phenotypic profiles, for identification of prevention efforts.

## Introduction

The initial stages of substance use disorder (SUD) often occur during adolescence, when youth begin to experiment with and, for some, regularly engage in substance use. Though problematic use patterns and full onset of SUD occur at a relatively young age (Lisdahl et al., 2021; Solmi et al., 2022), treatment for SUD is typically sought much later in life, if at all (de Girolamo et al., 2012; McGorry et al., 2011). Importantly, early prevention and intervention may stymy the progression to problematic use or SUD. Identification of biomarker predictors of SUD is then imperative for the development of early intervention efforts (Volkow et al., 2015). Accordingly, a large, longitudinal study called the Adolescent Brain Cognitive Development Study (ABCD) was launched by the U.S. National Institutes of Health in 2016, in part to identify biological and genetic vulnerabilities of children that predict later substance use (Volkow et al., 2018).

Previous studies demonstrate that SUD has a substantial genetic component, with heritability estimates ranging from 30 to 80% across alcohol, nicotine and other illicit substances (Agrawal & Lynskey, 2008; Schuckit, 2009; Urbanoski & Kelly, 2012). Genome-wide association studies (GWAS) and individual single nucleotide polymorphism (SNP) data can be used to quantify genetic liability by calculating a polygenic score (PGS). PGSs use an individual’s genetic makeup and an independent assessment of genetic associations to compute genetic risk for a given trait across their lifetime (Lewis & Vassos, 2020). It is suggested that PGS is most clinically useful for prevention efforts rather than intervention (Lambert et al., 2019).

PGS of substance use and SUD are demonstrated to increase predictive utility above that of family history alone (Schaefer et al., 2023; Wang et al., 2023). In a large cohort of Dutch adolescents, PGS were linked to any alcohol, nicotine, and cannabis use by age 16, but not frequency of use (Marceau et al., 2022). Further, our group previously showed a relationship between an opiate use disorder liability PGS and behavioral traits associated with substance use risk (specifically, behavioral inhibition) in an earlier wave of data from the ABCD Study (Loughnan et al., 2022), yet actual substance use behaviors were not investigated at that time. Most studies investigating PGSs and substance use examine late adolescence or young adulthood; identifying risk earlier in adolescence is important as early age of onset is one of the most robust predictors of negative clinical outcomes in youth who use substances (Lisdahl et al., 2013; Spear, 2015; Volkow et al., 2021).

Impulsivity is a multifaceted trait (Whiteside & Lynam, 2001) that is broadly linked to substance initiation in youth (Sonmez et al., 2024). Meta-analyses reveal unique patterns of relationships between impulsivity and substance use patterns, such as problematic cannabis and alcohol use differentially predicted by specific facets of impulsivity (Stautz & Cooper, 2013; VanderVeen et al., 2016). One study found sensation seeking (a facet of impulsivity) at age 13 mediates relationships between PGS and alcohol use problems by age 16 (Li et al., 2017). Prior work in the ABCD Study also reflects this; in a large machine learning-based analysis, sensation seeking was a robust predictor of substance use initiation by age 12 (Green et al., 2024). Therefore, investigating genetic risk for both impulsivity and substance use initiation in younger adolescents may inform how facets of impulsivity may translate to later problematic substance use.

In sum, studying substance use and its genetic contributions is crucial for identifying at-risk individuals and developing targeted prevention strategies, particularly during the vulnerable developmental period of adolescence. Here, using the PGS catalog, we generate PGSs across a range of substance use and related traits in over 11,000 individuals from the Adolescent Brain and Cognitive Development (ABCD) Study^®^. Youth were followed starting at ages 9-10, with data included here from the 2-Year and 4-Year follow up visits (through ages 13-14). We evaluate the performance of PGS to predict i) facets of impulsivity and ii) self-reported use of nicotine/tobacco, cannabis, alcohol, and caffeine, both across the entire sample and divided by age bands to probe developmental timing effects. We hypothesize that the relationships between PGS and impulsivity and substance use will strengthen as participants age.

## Methods

### ABCD Sample

The ABCD Study enrolled 11,878 youth ages 9-10 in 2016-2018 at 21 sites across the United States, and follows them with annual study visits (Jernigan et al., 2018). Epidemiologically guided, school-based probability sampling methods were used to recruit a sociodemographically diverse cohort (Garavan et al., 2018). Participants and their caregivers attend yearly appointments to their local study site to complete surveys and questionnaires on substance use (Lisdahl et al., 2018), mental and physical health (Barch et al., 2018; Barch et al., 2021), and culture and environment (Zucker et al., 2018). They also complete a neurocognitive battery (Luciana et al., 2018), contribute biosamples (Uban et al., 2018), and, every other year, complete a full magnetic resonance imaging protocol (Casey et al., 2018). Here, data from N= 11,389 participants at Baseline with full genetic data are included.

### Genomic Data

Genetic information was retrieved from saliva and blood sources and assayed using the Smokescreen™ Genotyping array (Baurley et al., 2016; Uban et al., 2018). Genetic principal components and genetic relatedness were calculated from ABCD genomic data using PC-Air (Conomos et al., 2015) and PC-Relate (Conomos et al., 2016), respectively, with default settings described elsewhere (Fan et al., 2023). To improve overlap with PGS posterior effects, the markers from the Smokescreen array were imputed using the TOPMED imputation server (Taliun et al., 2021). These imputed variants were fractional dosages that were converted to an integer number of alleles using a best guess threshold of 0.9. This resulted in 280,850,795 imputed variants aligned to genome build GRCh38. Using PLINK (v2) (Chang et al., 2015), we then restricted our target data to only autosomal variants with a minor appeal frequency of 1% (0.01) or greater which left us with 10,927,293 variants.

### Genomic Similarity to Continental Ancestry

As the predictive power of PGS are known to vary as genomic distance from the discovery GWAS increases (Ding et al., 2023), best practices dictate that PGS should be evaluated in populations that share similar genetic continental ancestry (Kachuri et al., 2024). To achieve this, we estimated similarity to continental ancestry groups for each individuals using SNPweights (Chen et al., 2013) and external genomic reference panels for African (AFR-like), East Asian (EAS-like), European (EUR-like) (Genomes Project et al., 2015), and Indigenous North and South American (AMR-like) (Reich et al., 2012) Ancestry Populations. ABCD subjects were separated either into continental ancestry groups if they had an inferred genetic ancestry of at least 80% consistent with a continental reference panel or as admixed ancestry (MIX-like) - i.e. those that were not 80% inferred genetic ancestry for any continental group.

Separating the samples into ancestral groups provided the following subsamples: EUR-like (n = 5225), AFR-like (n = 637), and a MIX-like (admixed) ancestry population (n = 2891). Table 1 and Supplementary Tables 1-2 show demographic features of each of these datasets. EAS-like and AMR-like ancestry groups were each less than 200 individuals and were considered too small to perform analysis in the current study. Graphical representations and analyses related to these ancestral groupings can be found in previous work done by Ahern and colleagues (Ahern et al., 2023). Due to the high representation of EUR-like PGS weights in training samples and the well documented issues of portability across ancestral groups (Ding et al., 2023), we present the primary results in this paper within the EUR-like cohort. In the supplementary material, we analyze AFR-like and MIX-like cohorts and present their findings.

**Table 1.** Sociodemographic information for EUR-like ancestry cohort.

|  | UPPS Analysis | Substance Use Analysis |
| --- | --- | --- |
| Total N at baseline | 5225 | 4958 |
| Age at baseline (SD) | 9.9 (0.62) | 9.9 (0.62) |
| Sex Female | 2424 | 2301 |
| Self-Declared Race/Ethnicity |  |  |
| White | 4593 | 4379 |
| Black | <10 | <10 |
| Hispanic | 397 | 357 |
| Asian | <10 | <10 |
| Other | 228 | 215 |

| Education |  |  |
| --- | --- | --- |
| Less than High School | 32 | 29 |
| GED | 1401 | 1304 |
| Associate's degree | 304 | 282 |
| Bachelor's | 1197 | 1133 |
| Master's | 1523 | 1470 |
| Graduate | 768 | 740 |

#### PGS Catalog

The Polygenic Score (PGS) Catalog is a curated, open-source, and constantly expanding database of PGS and detailed metadata. The goals of the PGS Catalog are to improve access to the data necessary to reproduce, compare, and independently use existing PGS (Lambert et al., 2021). PGS posterior effects were matched and applied to our ABCD target data using pgsc_calc (“The Polygenic Score Catalog Calculator (pgsc_calc),” 2023) maintained by the nf-core community, a group dedicated to designing collaborative, peer-reviewed, best-practice analysis pipelines with the goal of improving standardization and reproducibility in bioinformatics (Ewels et al., 2020). Scores were applied as long as they had a minimum variant overlap of at least 1% (0.01); otherwise, default parameters were used. We selected items from PGS Catalog that reported a trait related to substance use or a substance use disorder. This resulted in 116 PGS considered for subsequent analyses. One PGS (PGS000201: Problematic alcohol use) with an unclear definition of PGS calculation, hampering interpretation, was removed. Posterior weights from Saunders and colleagues (Saunders et al., 2022) had evidence of effect and reference allele mislabeling and the sign of the PGSs derived from this study (PGS003357, PGS003358, PGS003359, PGS003360, PGS003361, PGS003362, PGS003363, PGS003364, PGS003365, PGS003366, PGS003367, PGS003368, PGS003369, PGS003370, PGS003371, PGS003372, PGS003373, PGS003374, PGS003375, PGS003376) were flipped. Many PGS reported on the same named traits, some of these were also from the same study. In total out of the 116 PGS, there were only 37 unique reported traits. A list of PGS and phenotype definitions can be found in Supplementary Table 2. To avoid duplications of PGS on the same traits we employed a procedure to first match training and target ancestries, second to select PGS methods that employed continuous shrinkage techniques over lower performing pruning and thresholding techniques (Ahern et al., 2023), and finally, if multiple PGS still existed for a given trait, we selected the PGS with the largest training population. Specifically, the procedure was as follows:

1. To match training and target ancestries for analysis in the AFR-like cohort of ABCD (supplementary analysis), if a PGS AFR-like training sample was available this was selected. Otherwise, preference was given to PGS trained on a EUR-like training population.
2. When multiple methods were applied for a single trait, we selected a single PGS with the following preference: a) ‘BOLT-LMM’, b) ‘SbayesR’, c) ‘PRS-CS’, d) ‘Ldpred2 (bigsnpr)’, e) ‘Ldpred2-auto’, and f) ‘PRSice’. This reflects the broad finding of continuous shrinkage methods exhibiting superior performance compared to pruning and thresholding methods{Ni, 2021 #1807}.
3. If there still were multiple PGS, we selected the PGS with the largest training sample size.

### Impulsivity Measure

The UPPS-P Impulsive Behavior Scale measures five distinct facets of impulsivity (negative urgency, [lack of] premeditation, [lack of] perseverance, sensation seeking, and positive urgency) (Barch et al., 2018; VanderVeen et al., 2016). Youth participants completed the 20-item youth short version every other year of the ABCD Study, starting at Baseline, with 4 questions per subscale. This scale was modified from the full UPPS-P (Zapolski et al., 2010), adapted in conjunction with the adult short report (Lynam, 2013), to have a brief, developmentally appropriate measure of impulsivity from adolescence into adulthood. Data from Baseline, Year 2, and Year 4 are included here.

### Substance Use History

A full, detailed description of substance use reporting in ABCD is available elsewhere (Lisdahl et al., 2018). Briefly, trained study staff completed semi-structured interviews with youth participants regarding lifetime (at Baseline) and past-year substance use (subsequent annual visits). After querying what substances participants had heard of (Baseline and Year 2), participants were asked whether they had used each substance, including initial sipping of alcohol or puffing of nicotine or cannabis products. Participants were also oriented to reporting full standard units of each substance reported. Each drug class was individually queried. For analyses here, lifetime reporting of at least one standard unit of alcohol, nicotine/tobacco products, or cannabis was calculated. A binary endorsement for lifetime use by each drug, at each wave, was coded, using publicly available code (Sullivan et al., 2024). Notably, the vast majority of youth reporting nicotine product use endorsed vaping or use of electronic nicotine delivery systems (ENDS) (Sullivan et al., 2022).

For caffeine, participants self-reported average weekly number of total standard doses of caffeine beverages (8 oz for coffee or tea, an espresso shot, 12 oz soda, or 8 oz energy drink) for the past 6 months (baseline) or past month (subsequent annual visits). Any non-zero weekly endorsement was categorized as use of that caffeine product, this resulted in five caffeine measures related to weekly use of coffee, tea, espresso shots (including mochas and lattes), caffeinated sodas, and energy drinks.

### Statistical Methods

The ABCD 5.1 dataset (http://dx.doi.org/10.15154/z563-zd24) contains three major assessment time points with full behavioral and imaging study protocol: Baseline (9-10 years-old), Year 2 follow up (11-13 years-old), and Year 4 follow up (13-15 years-old). We conducted analysis in each wave of the study separately. As ABCD contains many related individuals including siblings and twins, to reduce bias resulting from this relatedness structure we performed analyses selecting singletons identified at the Baseline visit. The Year 4 follow up data collection was only partially complete with the 5.1 data release; as such this time point had roughly half the number of participants. We therefore conducted supplementary sensitivity analysis repeating all analyses restricted to those included in the 5.1 data release with data from the Year 4 time point to assess convergence of results when the sample was identical across all time points. Singletons were defined by generating a set of families where individuals were at least 35% genetically related to one another. This resulted in 8,753 families, across the three ancestry strata, from which singletons were derived for a total N of 8,753 across the present analyses.

Within each time point of the study, we separately fit linear regression models to predict a) each of the five subscales from the UPPS questionnaire, and b) each of the eight substance use related questions above (alcohol use, nicotine use, cannabis use, and the five caffeine categories). A separate model was fit using each PGS as an independent variable controlling for interview age, sex assigned at birth, highest parental education, study site, and top ten principal components of genetic ancestry. Parental education was coded as a five-point categorical variable indicating less than a high school degree, GED/high school degree, Associate’s degree, Bachelor’s degree, Master’s degree, or a higher graduate degree. We performed Bonferroni correction, indicated with p_bonf_, to account for multiple comparisons within each timepoint-analysis. For each timepoint of the UPPS analysis this amounted to 185 multiple tests, and for substance use analysis this was 296. Fixed effect variance explained by PGS on dependent variables was calculated from t-statistics and degrees of freedom (DF) as ΔR^2^=t^2^/(t^2^ + DF). The primary analyses of this paper are in the EUR-like cohort. We additionally performed all analyses in the AFR-like and MIX-like cohorts and present the concordance of these results with primary results in the supplementary results.

## Results

### Sociodemographic Descriptives

Table 1 presents sociodemographic of EUR-like ancestry cohort used in main analyses, with Supplementary Tables 1 and 2 reflecting sociodemographics of AFR-like and MIX-like ancestry cohorts.

### Primary Analyses

In-text reported results are limited to the five most statistically significant relationships by each year.

### UPPS-PGS Associations

Figure 1A displays associations between PGS and UPPS subscales across all timepoints with Supplementary Extended Data Table 1 containing a full summary of regression outputs.

**Figure 1.**
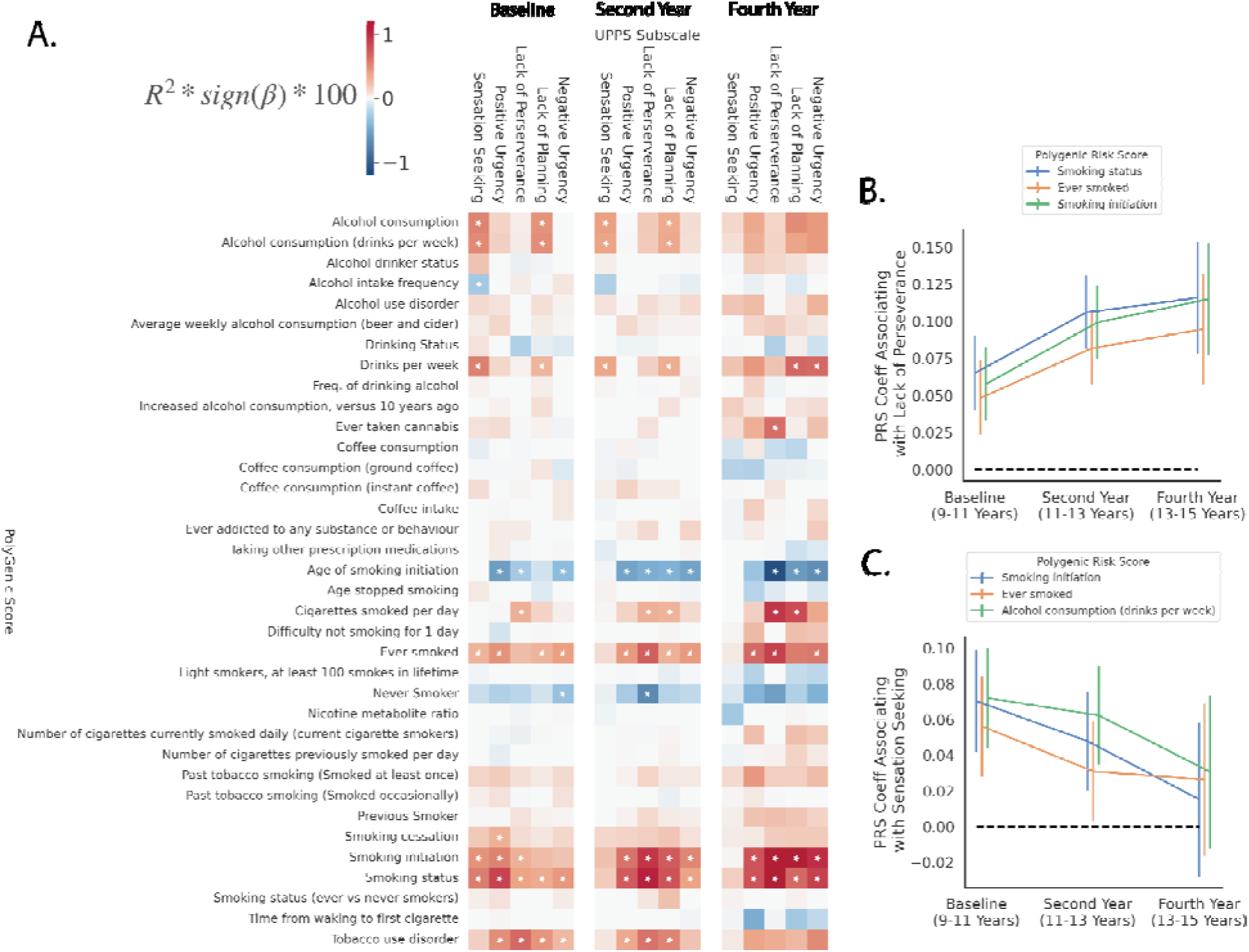
Associations between PGS and UPPS subscales within EUR-like samples across baseline, second year and fourth year time points. Panel A displays a heatmap with each element representing the variance explained as a percentage of the PGS with each UPPS subscale. Bonferroni significant p-values (corrected for tests within each time point) are indicated with stars. Panels B and C represent longitudinal changes shown as effect sizes (β: regression coefficients) and 95% confidence intervals of specific PGS with Lack of Perseverance, and Sensation Seeking subscales, respectively, across time points.

### Ages 9-11 (Baseline)

The most significant association at Baseline was between the PGS for Smoking Status (PGS002348) and Positive Urgency (ΔR^2^=0.011, p_bonf_=1.79×10^-12^). Tobacco use disorder (PGS002037) was significantly associated with Lack of Perseverance (ΔR^2^=0.0078, p_bonf_=3.49×10^-8^). Age of Smoking Initiation (for first use; PGS003364), Smoking Initiation (age of regular use; PGS003360), and Tobacco Use Disorder (PGS002037) all predicted Positive Urgency (ΔR^2^=0.0076, p_bonf_=5.51×10^-8^; ΔR^2^=0.0076, p_bonf_=6.27×10^-8^; ΔR^2^=0.0063, p_bonf_=1.51×10^-6^, respectively).

### Ages 11-13 (Year 2)

Current smoking status (PGS002348) was significantly associated with Lack of Perseverance (ΔR^2^=0.014, p_bonf_=6.78×10^-15^), Positive Urgency (ΔR^2^=0.0093, p_bonf_=9.05×10^-10^), and Lack of Planning (ΔR^2^=0.0085, p_bonf_=7.35×10^-9^). Smoking Initiation (age of regular use; PGS003360) and Smoking Status (never smoker; PGS001127) were significantly associated with Lack of Perseverance (ΔR^2^=0.012, p_bonf_=4.72×10^-13^; ΔR^2^=0.0085, p_bonf_=7.35×10^-9^, respectively).

### Ages 13-15 (Year 4)

Current smoking status (PGS002348) and Age of Smoking Initiation (for first use; PGS003364) were significantly associated with Lack of Perseverance (ΔR^2^=0.015, p_bonf_=4.07×10^-7^; ΔR^2^=0.013, p_bonf_=4.29×10^-6^). Smoking Initiation (age of regular use; PGS003360) predicted Lack of Perseverance (ΔR^2^=0.015, p_bonf_=4.12×10^-7^), Lack of Planning (ΔR^2^=0.014, p_bonf_=8.66×10^-7^), and Negative Urgency (ΔR^2^=0.011, p_bonf_=8.21×10^-5^).

### Associations By Time Point

We hypothesized that the association between PGS and impulsivity measures would strengthen as the sample aged and individuals, potentially indicative of increased risky behaviors. However, analyses revealed that this pattern was not consistently observed. We studied longitudinal patterns through inspecting regression coefficients as these are unbiased by differences in sample sizes between time points. As highlighted in Figure 1B, the Lack of Perseverance subscale showed an increasing effect with a number of PGS including: Smoking Status (PGS002348: Baseline β=0.065, p=3.43×10^-7^; Year 2 β=0.11, p=3.67×10^-17^; Year 4 β=0.12, p=2.20×10^-9^), Ever Smoked (PGS002126: Baseline β=0.048; p=1.38×10^-4^; Year 2 β=0.082, p=6.30×10^-11^; Year 4 β=0.095, p=8.70×10^-7^), and Smoking Initiation (PGS003360: Baseline β=0.058, p=5.94×10^-6^; Year 2 β=0.099, p=2.55×10^-15^; Year 4 β=0.11, p=2.22×10^-9^).

Conversely, as shown in Figure 1C, the effect between Sensation Seeking and a number of PGS diminished over time: Smoking initiation (PGS003360: Baseline β=0.065, p=5.17×10^-6^; Year 2 β=0.041, p=4.26×10^-3^; Year 4 β=0.012, p=0.057), Ever smoked (PGS002126: Baseline β=0.054; p=1.47×10^-4^, Year 2 β=0.028, p=0.044; Year 4 β=0.025, p=0.25), and Alcohol Consumption drinks per week (PGS000203: Baseline β=0.070, p=7.64×10^-7^; Year 2 β=0.062, p=1.26×10^-5^; Year 4 β=0.03, p=0.17). Although multiple nicotine related PGS (Age of smoking initiation PGS003364, Ever Smoked PGS002126, Smoking initiation PGS003360, Smoking status PGS002348, Tobacco Use Disorder PGS002037) were associated with all UPPS subscales at Baseline and Year 2, PGS of alcohol related traits (Alcohol Consumption PGS002839, Alcohol Consumption (drinks per week) PGS000203, and Drinks per Week PGS003376) only associated with Lack of Planning and Sensation Seeking traits at these same time points - see Figure 1A and Extended Data Table 1.

### Substance Use-PGS Associations

Figure 2A displays associations between PGS and substance use questions for each of the three waves of the study with supplementary Extended Data Table 1 containing a full summary of regression outputs.

### Ages 9-11 (Baseline)

Current smoking status (PGS002348) and Smoking Initiation (age of regular use; PGS003360) were significantly associated with regular Caffeinated Soda consumption (ΔR^2^=0.0065, p_bonf_=3.43×10^-6^; ΔR^2^=0.0050, p_bonf_=1.85×10^-4^). Current smoking status (PGS002348) and Smoking Initiation (age of regular use; PGS003360) were significantly associated with regular tea consumption (ΔR^2^=0.0061, p_bonf_=9.13×10^-6^; ΔR^2^=0.0036, p_bonf_=6.60×10^-4^).

### Ages 11-13 (Year 2)

Current Smoking Status (PGS002348), Smoking initiation (age of regular use; PGS003360), Smoking Status (never smoker; PGS001127), and Ever Smoked (yes/no; PGS002126) were all significantly associated with regular caffeine consumption (ΔR^2^=0.0066, p_bonf_=5.12×10^-6^; ΔR^2^=0.0063, p_bonf_=1.07×10^-5^; ΔR^2^=0.0054, p_bonf_=1.02×10^-4^, ΔR^2^=0.0044, p_bonf_=1.15×10^-3^)

### Ages 13-15 (Year 4)

Current Smoking status (PGS002348), Smoking Initiation (age of regular use; PGS003360), and Ever Smoked (PGS002126) all predicted nicotine/e-cigarette usage (ΔR^2^=0.012, p_bonf_=6.06×10^-5^; ΔR^2^=0.011, p_bonf_=8.46×10^-5^; ΔR^2^=0.0090, p_bonf_=1.50×10^-3^). Additionally, Smoking Initiation (age of regular use; PGS003360) predicted lifetime cannabis use (ΔR^2^=0.0094, p_bonf_=8.65×10^-4^) and Current Smoking status (PGS002348) predicted regular energy drink consumption (ΔR^2^=0.0093, p_bonf_=9.50×10^-4^).

### Associations By Time Point

As previously hypothesized for PGS-impulsivity associations, we expected that PGS and self-reported substance use associations would strengthen over time. However, once again, investigating longitudinal trends revealed this was not always the case. For example, several PGS associations with self-reported regular energy drink consumption (Figure 2B), including Smoking Status (PGS002348: Baseline β=0.088, p=0.41; Year 2 β=0.24, p=8.62×10^-4^; Year 4 β=0.29, p=3.21×10^-6^), Alcohol Consumption (drinks per week) (PGS000203: Baseline β=-0.032, p=0.75; Year 2 β=0.17, p=0.017; Year 4 β=0.17, p=5.38×10^-3^), and Smoking Initiation (PGS003360: Baseline β=0.094, p=0.37; Year 2 β=0.25, p=8.70×10^-4^; Year 4 β=0.23, p=1.92×10^-4^), exhibited a pattern of increasing association strength over time. In contrast, as depicted in Figure 2C, the effect between self-reported lifetime alcohol use and several PGS—including Alcohol Consumption (PGS002839: Baseline β=0.12, p=1.61×10^-4^; Year 2 β=0.11, p=2.65×10^-4^; Year 4 β=0.083, p = 0.066), Alcohol Consumption (drinks per week) (PGS000203: Baseline β=0.11, p=6.80×10^-4^; Year 2 β=0.10, p=9.17×10^-4^; Year 4 β=0.084, p=0.059), and Drinks per Week (PGS003376: Baseline β=0.12, p = 3.64×10^-4^; Year 2 β=0.11, p=3.16×10^-4^; Year 4 β=0.087, p=0.050)—displayed a declining trend. This pattern emerged despite an overall increase in self-reported alcohol use endorsement over time, as indicated by the right-hand y-axis in Figure 2C.

**Figure 2.**
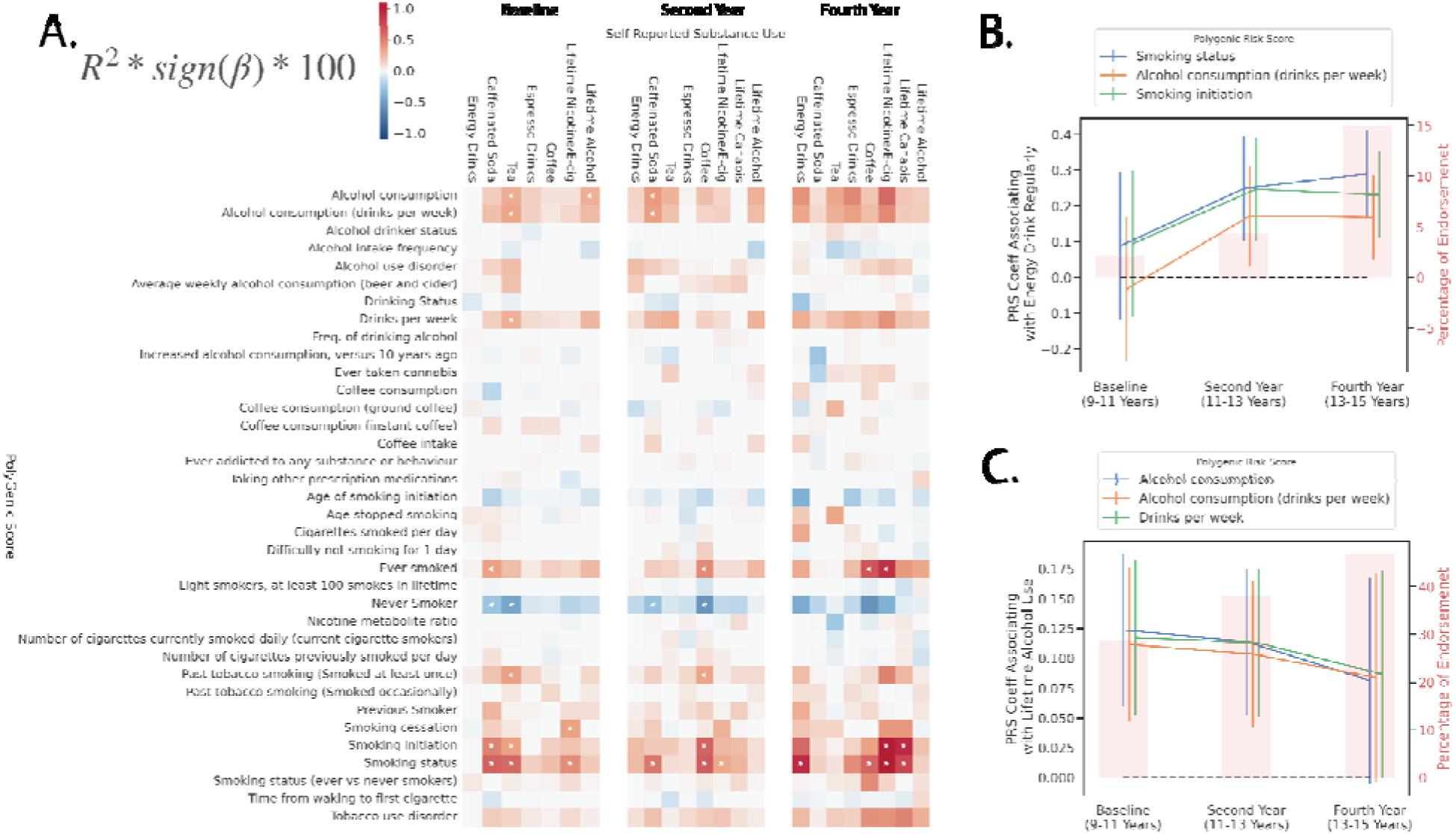
Associations between PGS and self-reported substance use within EUR-like samples across baseline, second year and fourth year time points. Panel A displays a heatmap with each element representing the variance explained as a percentage of the PGS with each substance use measure. Bonferroni significant p-values (corrected for tests within each time point) are indicated with stars. Panels B and C represent longitudinal changes shown as effect sizes (β, regression coefficients) and 95% confidence intervals of specific PGS with regular energy drink consumption, and lifetime alcohol use respectively, across time points. Right axis of panel B and C shows the percentage of endorsement at each time point for the relevant substance.

### Sensitivity Analysis of Singleton Selection

Since the main analyses selected singletons from the Baseline time point, but fewer individuals remained at the fourth-year time point, we conducted a sensitivity analysis to assess how selecting singletons from the smaller fourth-year sample might influence the results. Supplementary Figure 1 presents the findings of this analysis, demonstrating high concordance in results regardless of whether singletons were selected at Baseline or the Year 4 time point.

### Cross Ancestry Replication

Next, we evaluated the replication of results found with the EUR-like cohort in the MIX-like and AFR-like cohorts of ABCD (see Supplementary Tables 1 and 2 for demographics). For this analysis we observed modest replication in the MIX-like sample and no evidence of replication in the AFR-like sample (see Supplementary Figure 2). This is despite attempting to match AFR-like summary statistics with the AFR-like cohort where possible, see methods for details. As other researchers have noted we caution against interpreting results which are only significant in non EUR-like cohorts due to well documented issues of portability across ancestry groups{Ding, 2023 #1749}.

## Discussion

Here we assessed whether PGS predicted facets of impulsivity and substance use initiation over time in early to mid-adolescence, as understanding risk factors (including genetics) for at-risk youth is important for the development of substance use prevention and intervention efforts. Analyses across participants ages 9-15 revealed PGS of nicotine/tobacco smoking traits were associated with facets of impulsivity across time, particularly lack of perseverance. Similarly, overlapping PGS nicotine/tobacco traits were associated with early caffeine use behaviors and, by ages 13-15, lifetime cannabis use. Together, results indicate the dynamic relationship between genetic risk factors, behavioral (impulsivity) risk factors, and substance initiation across youth.

Impulsivity is established as a risk factor for substance use in adolescents, although analyses of genetic predictors of impulsivity outside of psychopathology (e.g., Attention Deficit Hyperactivity Disorder) are rare. Prior work in the ABCD Study found no alcohol use disorder PGS and UPPS-P relationships at Baseline (ages 9-10) (Su et al., 2022), which is partially replicated in the present analyses, though certain PGS traits (Number of Drinks Per Week; Problematic Alcohol Use) are associated with select facets of impulsivity (Sensation Seeking; Lack of Perseverance) and vary over time. It may be that impulsivity changes over time from a risk factor to a more developmentally normative trait. Specifically, most facets of impulsivity follow a linear decline from early to mid-adolescence, whereas sensation seeking increases through mid-adolescence and then declines (Harden & Tucker-Drob, 2011; Steinberg et al., 2008). It may also be that PGS, usually determined in adulthood, reflect different relationships than in adolescents; however, others previously showed adult PGS still predict adolescent externalizing symptoms and facets of impulsivity (Salvatore et al., 2015). Together, it does appear that PGS for nicotine/tobacco use offers a helpful phenotype for impulsivity and a target for prevention.

Interestingly, weekly caffeine use was largely linked with smoking polygenic risk traits across all three timepoints (Ages 9-15). These findings are consistent with the existing literature showing a link with early caffeine use and adolescent substance use initiation (Kristjansson et al., 2018). Caffeine and smoking, in particular, have long been linked (Kozlowski, 1976) with evidence in adult populations suggesting casual relationships between caffeine and smoking above broader genetic correlations (Chang et al., 2020). Our findings extend this literature showing links between caffeine and smoking as early as 9-years-old. Further, we found evidence suggesting increased associations between energy drinks specifically and PGS smoking traits as youth aged. This in part could be due to higher rates of energy drink consumption as youth get older (Miller et al., 2018), or increased underlying genetic risk with age. Indeed, early research shows that genetic influence over environmental risk increases on caffeine and nicotine relationships as individuals get older (Kendler et al., 2007). It may be that early caffeine use represents an important risk factor for later substance use. These findings provide support for the importance of investigating these relationships longitudinally in youth, given the current paucity of research in this area (Temple, 2019).

Tobacco related PGSs also predicted initial substance initiation over time in youth. We observed the largest number of associations for lifetime nicotine use across all time points, which was primarily characterized by youth self-reported use of electronic nicotine delivery systems (ENDS) or vaping e-cigarettes (Sullivan et al., 2022). Analyses of participants at later waves (i.e., ages 13-15) also revealed PGS-cannabis relationships that were not apparent at Baseline, when cannabis reporting was minimal. These findings broadly fit prior work of PGS predicting any substance use by mid-adolescence (Li et al., 2017; Marceau et al., 2022), and one study which found nicotine PGS predictive of use of other drug classes (Vink et al., 2014). While not necessarily prognostic, seeing these early relationships in youth who are often only experimenting with initial substance use confirms PGS as a risk marker.

Findings indicate differences in PGS prediction by age. For instance, smoking nicotine PGSs revealed a mostly linear increase in predicting Lack of Perseverance with increased age. As decreases in impulsivity are generally expected in typically developing youth (Harden & Tucker-Drob, 2011), this suggests a genetic liability to predict atypical adjustment to trait-based behaviors. There are also specific impulsivity profiles (Sensation Seeking and Lack of Planning) in alcohol-related PGS at ages 9-10 and 11-12 which may indicate a prodromal impulsivity risk profile that is specific to alcohol dependence in later life. For substance initiation prediction, smoking and alcohol PGS appear to largely increase in prediction of some domains as youth get older (e.g., energy drink use), whereas others (e.g., alcohol) PGS appear to decline in prediction of lifetime initiation of alcohol from ages 9-11 to ages 13-15. These divergent trends highlight the complexity of the relationship between genetic predisposition and substance use behaviors across developmental stages, suggesting that the influence of PGS may vary depending on the specific substance and the context of use. In addition, PGSs used here are from adults with often problematic levels of substance use. Prior data suggests PGS is not predictive of initiation, but escalation of nicotine use across adolescents (Belsky et al., 2013). Continued monitoring of PGS by actual substance use patterns in this cohort will be informative for identifying the most salient PGS markers and the most vulnerable age for assessment of risk to inform prevention efforts. Limitations include restricting our data to the PGS catalog, despite many other PGS exist.

In PGS catalog, the primary substance PGSs were related to alcohol, caffeine, and nicotine/tobacco products, with only two PGS scores for other substance use, limiting our ability to detect polygenic risk liability for other problematic substance use and impulsivity or substance initiation outcomes, as other groups have (Johnson et al., 2019). Most notably the PGS catalog does not contain results from most recent large scale GWAS conducted on cannabis use disorder (Levey et al., 2023), and instead only contains summary statistics from less targeted and less powered analysis in UK Biobank (Prive et al., 2022). Analyses here are cross-sectional, without accounting for within-subject outcome change in longitudinal analyses. Future longitudinal modeling of PGS and substance use outcomes, including escalation of substance use beyond initiation and including problematic use, is needed. Effect sizes were modest, and models did not include other environmental and trait factors which are linked to impulsivity and substance use. Future models should account for gene-by-environment associations, as the present models only explain a small portion of variance and these behavioral phenotypes are complex and multifaceted. Finally, PGS were derived from GWAS which were primarily conducted in EUR-like ancestry samples. Despite the diversity of the ABCD cohort and efforts to align training and target ancestries where possible, the predictive utility of these European-only PGS for non-European populations is limited which is underscored by our supplementary results. These results further underscore the pressing need for GWAS in non-European ancestry cohorts, as well as reaffirming the need for continued commitment in developing methods that can generalize findings across ancestry groups{Kachuri, 2024 #1758}{Ruan, 2022 #1809}.

In summary, analyses here indicate PGS for substance use and problems were predictive of impulsivity, a trait established as a predictor of substance use, and substance initiation itself across repeated cross-sectional analyses in youth 9-15. Polygenic risk relationships varied by age, with impulsivity and substance use relationships largely increasing over time. Significant associations between regular caffeine consumption and PGS, at a relatively illicit drug-naive age, may represent the detection of early addictive behaviors related to problematic substance use risk in later life. Results support PGS, in conjunction with larger phenotypic profiles, for identification of prevention efforts.

## Data Availability

The ABCD data repository grows and changes over time. The ABCD data used in this report came from http://dx.doi.org/10.15154/z563-zd24. DOIs can be found at http://dx.doi.org/10.15154/z563-zd24.

