## Supplementary Materials for "Predicting Genetic Risk for Impulsivity and Substance Use in Adolescence"

**Supplementary Material Index**:

**Supplementary Table 1** Demographics of MIX-like ancestry cohort used in supplementary analysis.

**Supplementary Table 2** Demographics of AFR-like ancestry cohort used in supplementary analysis.

**Supplementary Table 3**: Definitions of traits used for each PGS.

**Supplementary Figure 1:** Comparison of Singletons at Baseline and Year 4

**Supplementary Figure 2**: Cross-Ancestry PGS Comparisons

**Supplementary Table 1:** Demographics of MIX-like ancestry cohort used in supplementary analysis.

|  | UPPS Analysis | Substance Use Analysis |
| --- | --- | --- |
| Total N at baseline | 2891 | 2342 |
| Age at baseline (SD) | 9.9 (0.62) | 9.9 (0.62) |
| Sex Female | 1366 | 1106 |
| Self Declared Race/Ethnicity |  |  |
| White | 54 | 47 |
| Black | 704 | 561 |
| Hispanic | 1380 | 1044 |
| Asian | 62 | 54 |
| Other | 691 | 636 |
| Education |  |  |
| Less than High School | 251 | 147 |
| GED | 1354 | 1086 |
| Associates Degree | 319 | 253 |
| Bachelors | 425 | 356 |
| Masters | 375 | 344 |
| Graduate | 167 | 156 |

**Supplementary Table 2:** Demographics of AFR-like ancestry cohort used in supplementary analysis.

|  | UPPS Analysis | Substance Use Analysis |
| --- | --- | --- |
| Total N at baseline | 637 | 460 |
| Age at baseline (SD) | 9.9 (0.59) | 9.9 (0.59) |
| Sex Female | 318 | 230 |
| Self Declared Race/Ethnicity |  |  |
| White | 1 | 1 |
| Black | 611 | 437 |
| Hispanic | 5 | 4 |
| Asian | 0 | 0 |
| Other | 20 | 18 |
| Education |  |  |
| Less than High School | 42 | 25 |
| GED | 358 | 248 |
| Associates Degree | 82 | 61 |
| Bachelors | 64 | 48 |
| Masters | 73 | 63 |
| Graduate | 18 | 15 |

**Supplementary Table 3:** Definitions of traits used for each PGS.

| **PGS #** | **PGS Name** | **Definition** | **Scale** |
| --- | --- | --- | --- |
| PGS002839 | Alcohol consumption | Amount of alcohol drunk on a typical drinking day | Prefer not to answer/ 1 or 2/ 3 or 4/ 5 or 6/ 7, 8, or 9/ 10 or more |
| PGS000203  (Barr et al., 2020) | Alcohol consumption | Number of alcoholic drinks per week | Numeric value |
| PGS002115 | Alcohol drinker status | Alcohol drinker status | Prefer not to answer/ Never/ Previous/ Current |
| PGS002152 | Alcohol intake frequency | Alcohol intake frequency | Daily or almost daily/ Three or four times a week/ Once or twice a week/ One to three times a month/ Special occasions only/ Never/ Prefer not to answer |
| PGS002738    (Lai et al., 2022) | Alcohol use disorder | Scores from the problem subscale of the Alcohol Use Disorders Identification Test (AUDIT) | AUDIT Scores:  Range: 0-40  Low risk consumption: 1-7  Hazardous or harmful alcohol consumption: 8-14  Alcohol dependence (moderate-severe alcohol use disorder): 15 or more |
| PGS001088 | Average weekly alcohol consumption (beer and cider) | Average weekly beer plus cider intake | Do not know / Prefer not to answer/ Numeric value |
| PGS002913 | Drinking status | Alcohol drinker status | Never/ Past/ Current |
| PGS003376 | Drinks per week | Number of drinks per week | Numeric value |
| PGS001394 | Freq. of drinking alcohol | Frequency of drinking alcohol | Prefer not to answer / Never / Monthly or less / 2 to 4 times a month / 2 to 3 times a week/ 4 or more times a month |
| PGS001085 | Increased alcohol consumption, versus 10 years ago | Alcohol intake versus 10 years previously | Prefer not to answer/ Do not know/ Less nowadays/ About the same/ More nowadays |
| PGS002125 | Ever taken cannabis | Ever taken cannabis | Prefer not to answer/ No / Yes, 1-2 times / Yes, 3-10 times / Yes, 11 -100 times, Yes, more than 100 times |
| PGS001123 | Coffee consumption | Coffee consumed | Yes / No |
| PGS001124 | Coffee consumption (ground coffee) | Coffee type | Decaffeinated coffee (any type) / Instant coffee / Ground coffee (include espresso, filter etc) / Other type of coffee / Do not know / Prefer not to answer |
| PGS001125 | Coffee consumption (instant coffee) | Coffee type | Decaffeinated coffee (any type) / Instant coffee / Ground coffee (include espresso, filter etc) / Other type of coffee / Do not know / Prefer not to answer |
| PGS001126 | Coffee intake | Coffee intake | Do not know / Prefer not to answer/ Less than one/ Numeric value |
| PGS002124 | Ever addicted to any substance or behaviour | Ever addicted to any substance or behaviour | Prefer not to answer / Do not know / No / Yes |
| PGS001118 | Taking other prescription medications | Taking other prescription medications | Prefer not to answer / Do not know / No / Yes - you will be asked about this later by an interviewer |
| PGS003364 | Age of smoking initiation | The age at which the individual began smoking regularly | Years |
| PGS001374 | Age stopped smoking | Age stopped smoking | Do not know / Prefer not to answer / Year |
| PGS003368 | Cigarettes smoked per day | Amount smoked among current and former regular smokers was measured as cigarettes smoked per day | 1 = 1–5 per day; 2 = 6–15 per day; 3 = 16–25 per day; 4 = 26–35 per day; 5 = 36+ per day |
| PGS001388 | Difficulty not smoking for 1 day | Difficulty not smoking for 1 day | Prefer not to answer / Very difficult / Fairly difficult / Fairly easy / Very easy |
| PGS002126 | Ever smoked | Ever smoked | Yes / No |
| PGS001139 | Light smokers, at least 100 smokes in lifetime | Light smokers, at least 100 smokes in lifetime | Prefer not to answer / Do not know/ Yes / No |
| PGS001127 | Never Smoker | Smoking status | Prefer not to answer / Never / Previous / Current |
| PGS002233 | Nicotine metabolite ratio | The ratio of two stable nicotine metabolites: 3-hydroxycotinine to cotinine measured in the blood of current smokers | Numeric value |
| PGS001515 | Number of cigarettes currently smoked daily | Number of cigarettes currently smoked daily (current cigarette smokers) | Do not know / Prefer not to answer/ Less than one/ Numeric value |
| PGS001130 | Number of cigarettes previously smoked per day | Number of cigarettes previously smoked daily | Do not know / Less than one/ Numeric value |
| PGS001046 | Past tobacco smoking (Smoked at least once) | Past tobacco smoking | Prefer not to answer / I have never smoked / Just tried once or twice / smoked occasionally / Smoked on most or all days |
| PGS001047 | Past tobacco smoking (Smoked occasionally) | Past tobacco smoking | Prefer not to answer / I have never smoked / Just tried once or twice / smoked occasionally / Smoked on most or all days |
| PGS001128 | Previous smoker | Smoking status | Prefer not to answer / Never / Previous / Current |
| PGS003372 | Smoking cessation | Smoking cessation[9] (SmkCes; n = 1,400,535) contrasted current versus former smokers | Current smokers = 2  Former smokers = 1  Never smokers = missing |
| PGS003360 | Smoking initiation | Measures of onset included whether an individual ever smoked regularly and the age at which the individual began smoking regularly | Age (in years) at which an individual started smoking cigarettes regularly |
| PGS002348 | Smoking status | Smoking status | Prefer not to answer / Never / Previous / Current |
| PGS001129 | Smoking status (ever vs never smokers) | Tobacco smoking | Prefer not to answer / Never smoked / Ex- smoker / Occasionally / Smokes on most or all days |
| PGS001532 | Time from waking to first cigarette | Time from waking to first cigarette | Prefer not to answer / Do not know / Longer than 2 hours /Between 1 and 2 hours / Between 30 minutes - 1 hour / Between 5-15 minutes / Less than 5 minutes |
| PGS002037 | Tobacco Use Disorder | Identified using phecode 318.0 which corresponds to ICD codes F17.0, F17.1, F17.2, F17.3, F17.4, F17.9, Z72.0, 305.1, 305.10, 305.11, 305.12, 305.13, 649.0, 649.00, 649.01, 649.02, 649.03, 649.04 and V15.82 | Presence of tobacco use disorder diagnosis in electronic health records |

**Supplementary Figure 1:** Comparison of Singletons at Baseline and Year 4

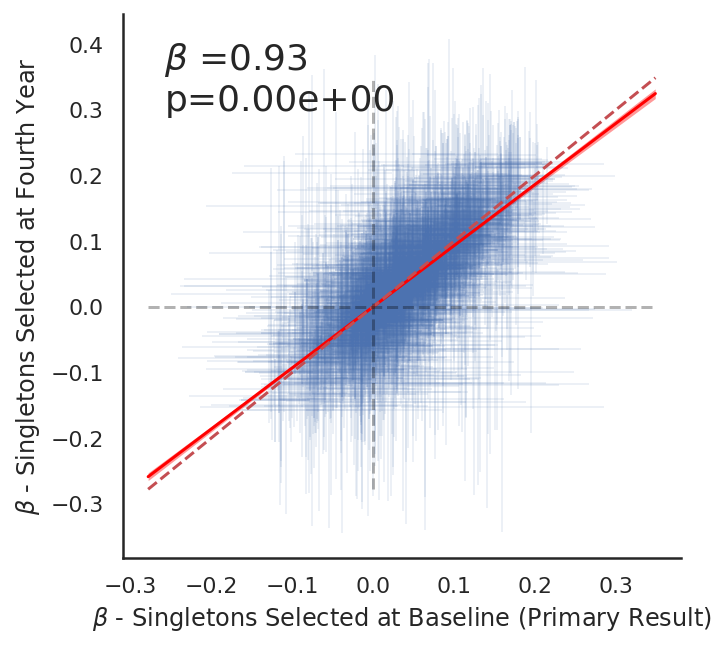

Notes: Sensitivity analysis to investigate the effect of selecting singletons from baseline cohort (x-axis) resulting in 5,225 individuals or selecting singletons from fourth year (y-axis) resulting in 2,423 individuals. Each beta coefficient represents a PRS and behavior association (UPPS or Self Reported Substance Use) with the EUR-like ancestry cohort used in the primary results.

**Supplementary Figure 2**: Cross-Ancestry PGS Comparisons

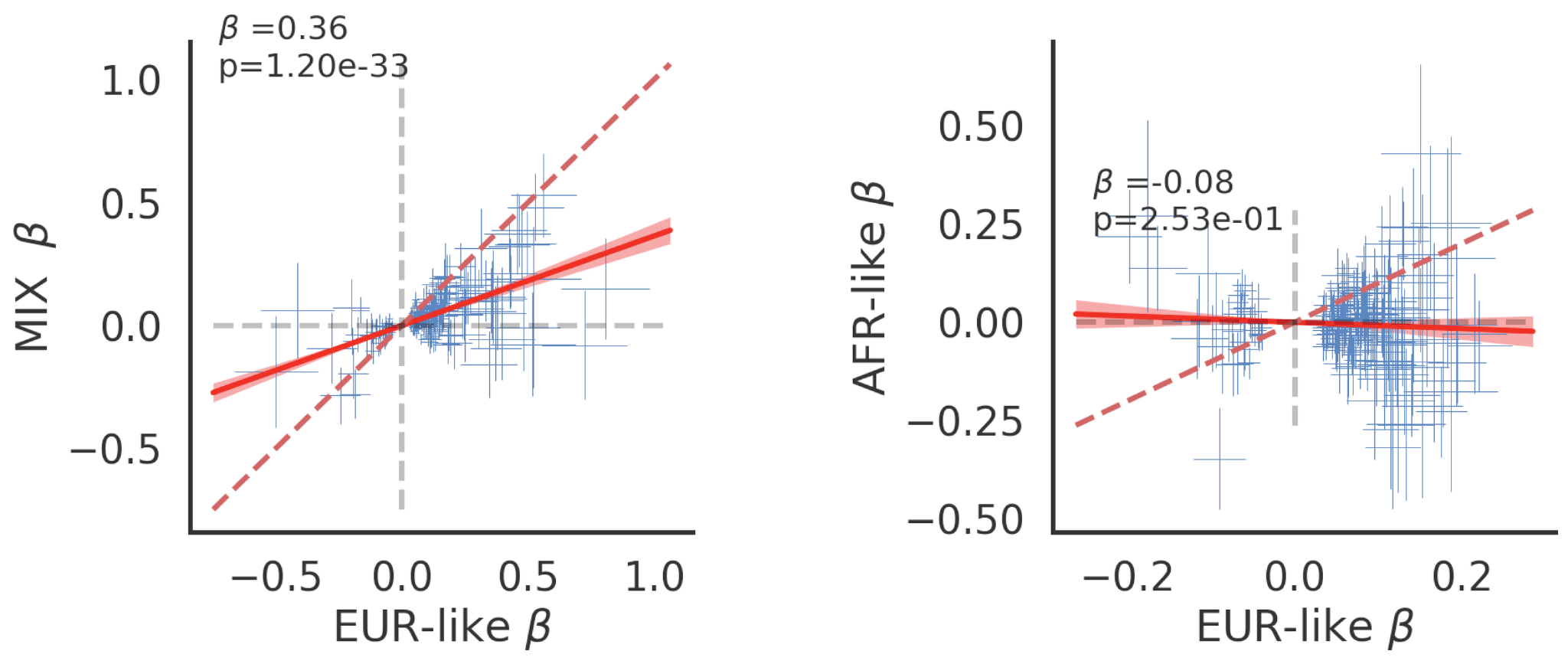

Notes: Cross ancestry application left panel for MIX cohort, right panel for AFR-like cohort. Each point represents a significant PGS vs behavior association that was significant in the EUR-like (main result) cohort. For substance use associations a minimum number of 20 people endorsing the question was required for both EUR-like and cross ancestry cohort to be included in the plot. Modest replication is observed in the MIX-like cohort and no replication is observed in the AFR-like cohort.
